# Correlates of DHS-defined infecund/menopausal status among Nigerian women aged 45-49: Evidence from the 2024 Nigeria Demographic and Health Survey

**DOI:** 10.64898/2026.06.04.26354907

**Authors:** Olubusayo Bankole Ogunsemoyin, Aboluwaji Daniel Ayinmoro

## Abstract

**Introduction:** Women aged 45-49 occupy a heterogeneous late-reproductive-life stage, but population research often treats them as a uniform group. This study examined correlates of Demographic and Health Survey (DHS)-defined infecund/menopausal status among Nigerian women aged 45-49.

**Methods:** This cross-sectional secondary analysis used the 2024 Nigeria Demographic and Health Survey Women Recode dataset. Weighted descriptive statistics summarised reproductive exposure status among 3,237 women. Out of these, 3,110 women classified as either fecund or infecund/menopausal were subjected to Survey-adjusted Chi-square tests and Binary Logistic regression at p<0.05, where pregnant and postpartum amenorrhoeic women were excluded.

**Results:** More than half of women were classified as infecund/menopausal (54.1%), while 41.5% were fecund; 3.2% were postpartum amenorrhoeic, and 1.3% were pregnant. Findings indicated that currently married/cohabiting women (AOR=4.87; 95% CI: 2.24-10.56) and formerly married women (AOR=8.30; 95% CI: 3.69-18.66) had higher odds of infecund/menopausal classification than women never in a union. Secondary education, higher education, middle-to-richest wealth quintiles, and five or more children ever born were associated with lower odds, while Northern minority ethnicity was associated with higher odds. Adding the current contraceptive method attenuated several education, wealth and parity associations; modern-method and traditional-method users had markedly lower odds than non-users.

**Conclusion:** Late-reproductive-life exposure status among Nigerian women aged 45-49 is socially patterned, with union status showing the most stable association. DHS-defined infecund/menopausal status is a demographic exposure category rather than clinically confirmed menopause. It is therefore concluded that the cross-sectional associations should not be interpreted causally.

## Introduction

The years just before the 50 mark a distinct stage in women’s reproductive lives. Some women are still conceiving, some are pregnant or recently postpartum, and others are in peri- or postmenopause [1,2]. Menopause timing varies widely, and although fecundity declines, it is not absent. Studies have also shown that pregnancy risks are substantially higher at ≥45 [2-5]. For demographers, clinicians, and programmes, this means that late-reproductive-age women cannot be treated as uniformly non-reproductive; careful fertility measurement and tailored contraceptive counselling up to the mid-50s are warranted [6-9].

Population surveys do not provide clinical confirmation of menopause for all respondents. Instead, the DHS reproductive exposure variable (V623) classifies women as fecund, pregnant, postpartum amenorrhoeic, or infecund/menopausal [10]. For women aged 45-49, this measure permits analysis of late-reproductive-life exposure status, provided its demographic meaning is not conflated with clinically confirmed menopause or infertility. The 2024 Nigeria Demographic and Health Survey (NDHS) provides a national data source for examining this classification in the Nigerian population [11].

Evidence on menopausal timing indicates that reproductive ageing is socially patterned. International reviews have associated lower socioeconomic position, lower education and selected reproductive factors with earlier age at natural menopause, although the direction and strength of associations vary across populations [12,13]. Nigerian studies have documented menopause-related experiences and factors associated with the attainment or timing of menopause in specific communities and clinical populations [14-16]. However, nationally representative evidence on DHS-defined infecund/menopausal status among Nigerian women aged 45-49 remains limited.

This gap matters because treating women aged 45-49 as a homogeneous post-reproductive group may obscure variation in continued exposure to pregnancy and the social distribution of late-reproductive-life status. The present study, therefore, examined correlates of DHS-defined infecund/menopausal status among Nigerian women aged 45-49 using the 2024 NDHS. Specifically, it described reproductive exposure status; examined bivariate variation by selected social, demographic and reproductive characteristics; and estimated survey-adjusted associations with infecund/menopausal classification.

## Theoretical framework

This study was guided by life-course and social determinants perspectives. A life-course approach conceptualises reproductive status at ages 45-49 as embedded in accumulated marital, reproductive and social trajectories rather than reducible to chronological age alone [17]. Union history and parity are therefore interpreted as markers of prior reproductive exposure and linked-life processes, not as direct biological causes of menopause.

The social determinants perspective directs attention to patterned inequalities in education, material resources, livelihood circumstances and sociocultural context that may distinguish women’s reproductive-life experiences [18]. In this analysis, education, wealth, residence, work status, religion and ethnicity are examined as social correlates of DHS-defined exposure status. The framework supports the assessment of population patterning while cautioning against causal or clinical inference from cross-sectional survey classifications.

## Materials and methods

### Study design and data source

This study was a cross-sectional secondary analysis of the 2024 Nigeria Demographic and Health Survey (NDHS) Women Recode dataset. The NDHS was implemented by the Federal Ministry of Health and Social Welfare of Nigeria, the National Population Commission, and ICF, using a nationally representative, stratified, two-stage sampling design [11]. The Women Recode dataset was selected because reproductive exposure status and the explanatory variables used in this study are measured at the individual woman level. The dataset was accessed for research purposes on 4 May 2026, following approval through the DHS Program data access procedure.

The analytic population was restricted to women aged 45-49 years. This restriction produced 3,237 women for the descriptive analysis. For binary analyses, women classified as pregnant or postpartum amenorrhoeic were excluded because the substantive contrast of interest was between women classified as fecund and those classified as infecund/menopausal. The binary analytic sample comprised 3,110 women, with no missing data on variables retained in either adjusted specification.

### Outcome variable

The dependent variable was DHS reproductive exposure status (V623). In descriptive analysis, the four original categories were presented: fecund, pregnant, postpartum amenorrhoeic, and infecund/menopausal. For bivariate and logistic regression analyses, the outcome was coded as 0 = fecund and 1 = infecund/menopausal. This operationalisation addresses demographic reproductive exposure status; it does not diagnose natural menopause, infertility or pathological amenorrhoea.

### Explanatory variables

Explanatory variables were selected on conceptual grounds from the life-course and social determinants framework. They included place of residence (urban, rural), education (no education, primary, secondary, higher), wealth quintile (poorest, poorer, middle, richer, richest), union status (never in union, currently married/cohabiting, formerly married), children ever born (0, 1-4, 5+), religion (Christianity, Islam, traditional/other), ethnicity (Hausa/Fulani, Yoruba, Igbo/Ibo, Northern minority, Southern minority, unclassified), and working status (no, yes). Age at first cohabitation or marriage was examined in the unadjusted analysis but excluded from the adjusted models because its never-in-union/not-applicable category structurally overlaps with current union status.

The current contraceptive method was coded as ‘not using’, ‘modern method’, or ‘traditional method’. It was excluded from the primary adjusted model because it was measured at the same survey point as reproductive exposure status and is conceptually close to continuing pregnancy exposure. It was introduced in a second model solely to examine the sensitivity of adjusted associations to this measurement-proximal correlate.

### Statistical analysis

Weighted percentages described reproductive exposure status and background characteristics. Weighted cross-tabulations with design-adjusted chi-square tests assessed bivariate variation in the binary outcome. Survey-adjusted binary logistic regression estimated unadjusted odds ratios (UORs) and adjusted odds ratios (AORs) with 95% confidence intervals (CIs). Model 1 was the primary adjusted specification and excluded the current contraceptive method; Model 2 added this variable as a sensitivity specification. Model 1 and Model 2 were estimated on the same binary analytic sample.

All analyses accounted for the NDHS complex sample design: women’s sample weights were applied as V005/1,000,000, primary sampling units were specified in V021, and strata were specified in V023. Analyses were conducted using IBM SPSS Statistics version 29. Given the cross-sectional design and the composite nature of the outcome, results are presented as associations and not causal effects.

### Ethical considerations

This study used de-identified secondary data from the 2024 NDHS. Ethical approval and informed consent procedures for the original survey are reported in the NDHS final report [11]. The authors did not participate in the original data collection, had no direct contact with survey participants, and did not have access to information that could identify individual participants during or after data collection. The dataset released to secondary users was anonymised before access. Therefore, separate institutional ethical approval was not sought for this secondary analysis. The authors obtained permission to use the dataset through The DHS Program data access procedure and complied with the approved data-use conditions.

## Results

### Reproductive exposure status and background characteristics

Table 1 presents the weighted profile of women aged 45-49. More than half were classified as infecund/menopausal (54.1%), while 41.5% remained classified as fecund. Only small proportions were postpartum amenorrhoeic (3.2%) or pregnant (1.3%). The women were almost evenly distributed between urban and rural residences, while about four in five were working, and more than four in five were currently married or cohabiting. Almost two-thirds had five or more children ever born, and about four in five were not using a contraceptive method at the time of the survey.

**Table 1.** Weighted descriptive profile of Nigerian women aged 45-49, NDHS 2024.

| Characteristic | Category | Unweighted n | Weighted % |
| --- | --- | --- | --- |
| Reproductive exposure status | Fecund | 1,322 | 41.5 |
|  | Pregnant | 42 | 1.3 |
|  | Postpartum amenorrhoeic | 85 | 3.2 |
|  | Infecund/menopausal | 1,788 | 54.1 |
| Place of residence | Urban | 1,611 | 50.6 |
|  | Rural | 1,626 | 49.4 |
| Ethnicity | Hausa/Fulani | 840 | 32.9 |
|  | Yoruba | 550 | 18.2 |
|  | Igbo/Ibo | 618 | 14.2 |
|  | Northern minority | 700 | 20.9 |
|  | Southern minority | 466 | 12.2 |
|  | Unclassified | 63 | 1.7 |
| Education | No education | 1,210 | 41.2 |
|  | Primary | 694 | 20.0 |
|  | Secondary | 881 | 25.6 |
|  | Higher | 452 | 13.3 |
| Religion | Christianity | 1,879 | 51.1 |
|  | Islam | 1,324 | 48.1 |
|  | Traditional/other | 34 | 0.8 |
| Wealth quintile | Poorest | 551 | 16.8 |
|  | Poorer | 493 | 16.6 |
|  | Middle | 655 | 19.4 |
|  | Richer | 732 | 20.9 |
|  | Richest | 806 | 26.3 |
| Union status | Never in union | 76 | 2.0 |
|  | Currently married/cohabiting | 2,652 | 83.2 |
|  | Formerly married | 509 | 14.8 |
| Children ever born | 0 | 119 | 3.3 |
|  | 1-4 | 1,132 | 34.0 |
|  | 5+ | 1,986 | 62.7 |
| Current contraceptive method | Not using | 2,604 | 81.4 |
|  | Modern method | 445 | 12.7 |
|  | Traditional method | 188 | 5.9 |
| Working status | No | 563 | 19.0 |
|  | Yes | 2,674 | 80.9 |

Source: Computed from the 2024 NDHS Women Recode dataset. Notes: Analysis is restricted to women aged 45-49 years ( $n = 3,237$ ). Weighted percentages use $V005/1,000,000$ and may not total exactly 100.0 because of rounding.

### Bivariate analysis of infecund/menopausal status women by socio-demographic characteristics

Table 2 shows weighted row distributions for women classified as either fecund or infecund/menopausal. Infecund/menopausal classification differed significantly by residence, ethnicity, education, wealth quintile, union status, current contraceptive method and working status. It was more prevalent among rural than urban women, among women with no education than those with higher education, and among women in the poorest rather than the richest quintile.

**Table 2.** Bivariate analysis of DHS-defined infecund/menopausal status among Nigerian women aged 45-49.

| Characteristic | Category | Fecund % | Infecund/menopausal % | Design-adjusted chi-square | p-value |
| --- | --- | --- | --- | --- | --- |
| **Place of residence:** |  |  |  | 19.29 | <0.001 |
|  | Urban | 48.1 | 51.9 |  |  |
|  | Rural | 38.5 | 61.5 |  |  |
| **Ethnicity:** |  |  |  | 25.51 | <0.001 |
|  | Hausa/Fulani | 43.3 | 56.7 |  |  |
|  | Igbo/Ibo | 41.5 | 58.5 |  |  |
|  | Yoruba | 53.2 | 46.8 |  |  |
|  | Northern minority | 37.5 | 62.5 |  |  |
|  | Southern minority | 41.0 | 59.0 |  |  |
|  | Unclassified | 44.2 | 55.8 |  |  |
| **Education:** |  |  |  | 30.02 | <0.001 |
|  | No education | 38.1 | 61.9 |  |  |
|  | Primary | 40.8 | 59.2 |  |  |
|  | Secondary | 47.8 | 52.2 |  |  |
|  | Higher | 54.4 | 45.6 |  |  |
| **Religion:** |  |  |  | 0.47 | 0.792 |
|  | Christianity | 43.2 | 56.8 |  |  |
|  | Islam | 43.7 | 56.3 |  |  |
|  | Traditional/other | 35.9 | 64.1 |  |  |
| **Wealth quintile:** |  |  |  | 36.27 | <0.001 |
|  | Poorest | 32.4 | 67.6 |  |  |
|  | Poorer | 38.1 | 61.9 |  |  |
|  | Middle | 41.7 | 58.3 |  |  |
|  | Richer | 47.2 | 52.8 |  |  |
|  | Richest | 51.3 | 48.7 |  |  |
| **Union status:** |  |  |  | 37.26 | <0.001 |
|  | Never in union | 74.5 | 25.5 |  |  |
|  | Currently married/cohabiting | 44.7 | 55.3 |  |  |
|  | Formerly married | 31.9 | 68.1 |  |  |
| **Children ever born:** |  |  |  | 0.64 | 0.725 |
|  | 0 | 41.3 | 58.7 |  |  |
|  | 1-4 | 42.4 | 57.6 |  |  |
|  | 5+ | 44.1 | 55.9 |  |  |
| **Current contraceptive method:** |  |  |  | 268.18 | <0.001 |
|  | Not using | 32.6 | 67.4 |  |  |
|  | Modern method | 87.4 | 12.6 |  |  |
|  | Traditional method | 91.5 | 8.5 |  |  |
| **Working status:** |  |  |  | 33.55 | <0.001 |
|  | No | 36.6 | 63.4 |  |  |
|  | Yes | 44.9 | 55.1 |  |  |

Source: Computed from the 2024 NDHS Women Recode dataset. Notes: Analysis is restricted to women classified as either fecund or infecund/menopausal (n = 3,110); pregnant and postpartum amenorrhoeic women were excluded. Percentages are weighted row percentages. Tests account for sample weights, primary sampling units and strata.

Formerly, married women had the highest proportion classified as infecund/menopausal, whereas women never in a union had the lowest proportion. No statistically significant differences were observed by religion or the number of children ever born.

### Multivariate analysis of infecund/menopausal status women using a logistic regression model

Table 3 presents the unadjusted estimates and Model 1, the primary adjusted model excluding the current contraceptive method. In Model 1, the bivariate association with residence was no longer statistically significant. Compared with women with no education, those with secondary or higher education had lower adjusted odds of an infecund/menopausal classification. Women in the middle, richer and richest wealth quintiles also had lower adjusted odds than those in the poorest quintile.

**Table 3.** Unadjusted and Model 1 survey-adjusted odds ratios for DHS-defined infecund/menopausal status among Nigerian women aged 45-49.

| Variable | UOR (95% CI) | p-value | Model 1 AOR (95% CI) | p-value |
| --- | --- | --- | --- | --- |
| Residence: Urban (RC) | 1.00 |  | 1.00 |  |
| Rural | 1.48 (1.24-1.76) | <0.001 | 1.08 (0.87-1.35) | 0.471 |
| Education: No education (RC) | 1.00 |  | 1.00 |  |
| Primary | 0.89 (0.70-1.13) | 0.337 | 0.91 (0.68-1.22) | 0.519 |
| Secondary | 0.67 (0.55-0.83) | <0.001 | 0.71 (0.53-0.95) | 0.023 |
| Higher | 0.52 (0.39-0.68) | <0.001 | 0.55 (0.38-0.79) | 0.001 |
| Wealth quintile: Poorest (RC) | 1.00 |  | 1.00 |  |
| Poorer | 0.78 (0.55-1.10) | 0.158 | 0.74 (0.52-1.06) | 0.101 |
| Middle | 0.67 (0.50-0.90) | 0.008 | 0.67 (0.47-0.94) | 0.020 |
| Richer | 0.54 (0.40-0.72) | <0.001 | 0.55 (0.38-0.80) | 0.002 |
| Richest | 0.46 (0.34-0.61) | <0.001 | 0.52 (0.35-0.78) | 0.002 |
| Union status: Never in a union (RC) | 1.00 |  | 1.00 |  |
| Currently married/cohabiting | 3.61 (1.96-6.64) | <0.001 | 4.87 (2.24-10.56) | <0.001 |
| Formerly married | 6.23 (3.24-11.98) | <0.001 | 8.30 (3.69-18.66) | <0.001 |
| Age at first cohabitation: <18 (RC) | 1.00 |  | - | - |
| 18-19 | 0.72 (0.55-0.95) | 0.019 | - | - |
| 20-24 | 0.56 (0.44-0.70) | <0.001 | - | - |
| 25+ | 0.58 (0.46-0.74) | <0.001 | - | - |
| Never in union/not applicable | 0.18 (0.10-0.34) | <0.001 | - | - |
| Children ever born: 0 (RC) | 1.00 |  | 1.00 |  |
| 1-4 | 0.95 (0.61-1.50) | 0.841 | 0.68 (0.38-1.22) | 0.194 |
| 5+ | 0.89 (0.57-1.39) | 0.616 | 0.45 (0.25-0.81) | 0.007 |
| Religion: Christianity (RC) | 1.00 |  | 1.00 |  |
| Islam | 0.98 (0.82-1.17) | 0.827 | 0.91 (0.68-1.22) | 0.546 |
| Traditional/other | 1.36 (0.52-3.53) | 0.528 | 0.97 (0.39-2.42) | 0.941 |
| Ethnicity: Hausa/Fulani (RC) | 1.00 |  | 1.00 |  |
| Yoruba | 0.67 (0.52-0.87) | 0.003 | 0.95 (0.65-1.39) | 0.796 |
| Igbo/Ibo | 1.08 (0.81-1.43) | 0.618 | 1.46 (0.96-2.23) | 0.078 |
| Northern minority | 1.27 (1.00-1.62) | 0.049 | 1.36 (1.01-1.83) | 0.040 |
| Southern minority | 1.10 (0.83-1.47) | 0.513 | 1.44 (0.94-2.21) | 0.091 |
| Unclassified | 0.96 (0.50-1.87) | 0.912 | 1.00 (0.49-2.04) | 0.995 |
| Current contraceptive method: Not using (RC) | 1.00 |  | - | - |
| Modern method | 0.07 (0.05-0.10) | <0.001 | - | - |
| Traditional method | 0.04 (0.02-0.09) | <0.001 | - | - |
| Working status: No (RC) | 1.00 |  | 1.00 |  |
| Yes | 0.71 (0.55-0.90) | 0.005 | 0.81 (0.63-1.04) | 0.099 |

Source: Computed from the 2024 NDHS Women Recode dataset. Notes: Outcome is coded 1 = infecund/menopausal and 0 = fecund. Pregnant and postpartum amenorrhoeic women were excluded. Model 1 adjusts for residence, education, wealth quintile, union status, children ever born, religion, ethnicity and working status. Age at first cohabitation was not included in Model 1 because it structurally overlaps with union status. RC = reference category; UOR = unadjusted odds ratio; AOR = adjusted odds ratio; CI = confidence interval.

Union status showed the strongest association in Model 1. Relative to women never in a union, currently married/cohabiting women had nearly five times the adjusted odds of infecund/menopausal classification. In comparison, formerly married women had more than eight times the adjusted odds. Women with five or more children ever born had lower adjusted odds than women with no children, and Northern minority women had higher adjusted odds than Hausa/Fulani women. Religion and working status were not significantly associated after adjustment.

Table 4 compares Model 1 with Model 2, which includes the current contraceptive method. The associations for current and former union status persisted across both adjusted models. In contrast, associations for secondary and higher education, the middle-to-richest wealth quintiles, and five or more children ever born were attenuated. They no longer met the 5% significance threshold in Model 2. Igbo/Ibo ethnicity was significant only after the inclusion of the current contraceptive method, whereas Northern minority ethnicity remained significant in both specifications.

**Table 4.** Sensitivity comparison of adjusted models for DHS-defined infecund/menopausal status among Nigerian women aged 45-49.

| Variable | Model 1 AOR (95% CI) | p-value | Model 2 AOR (95% CI) | p-value |
| --- | --- | --- | --- | --- |
| Residence: Urban (RC) | 1.00 |  | 1.00 |  |
| Rural | 1.08 (0.87-1.35) | 0.471 | 1.03 (0.80-1.33) | 0.794 |
| Education: No education (RC) | 1.00 |  | 1.00 |  |
| Primary | 0.91 (0.68-1.22) | 0.519 | 0.98 (0.72-1.35) | 0.919 |
| Secondary | 0.71 (0.53-0.95) | 0.023 | 0.84 (0.61-1.17) | 0.314 |
| Higher | 0.55 (0.38-0.79) | 0.001 | 0.70 (0.46-1.07) | 0.103 |
| Wealth quintile: Poorest (RC) | 1.00 |  | 1.00 |  |
| Poorer | 0.74 (0.52-1.06) | 0.101 | 0.79 (0.55-1.13) | 0.198 |
| Middle | 0.67 (0.47-0.94) | 0.020 | 0.72 (0.50-1.04) | 0.078 |
| Richer | 0.55 (0.38-0.80) | 0.002 | 0.68 (0.45-1.03) | 0.066 |
| Richest | 0.52 (0.35-0.78) | 0.002 | 0.68 (0.43-1.07) | 0.097 |
| Union status: Never in a union (RC) | 1.00 |  | 1.00 |  |
| Currently married/cohabiting | 4.87 (2.24-10.56) | <0.001 | 5.28 (2.20-12.65) | <0.001 |
| Formerly married | 8.30 (3.69-18.66) | <0.001 | 6.28 (2.57-15.37) | <0.001 |
| Children ever born: 0 (RC) | 1.00 |  | 1.00 |  |
| 1-4 | 0.68 (0.38-1.22) | 0.194 | 1.07 (0.60-1.93) | 0.810 |
| 5+ | 0.45 (0.25-0.81) | 0.007 | 0.75 (0.42-1.34) | 0.331 |
| Religion: Christianity (RC) | 1.00 |  | 1.00 |  |
| Islam | 0.91 (0.68-1.22) | 0.546 | 0.70 (0.49-1.00) | 0.048 |
| Traditional/other | 0.97 (0.39-2.42) | 0.941 | 1.25 (0.42-3.70) | 0.690 |
| Ethnicity: Hausa/Fulani (RC) | 1.00 |  | 1.00 |  |
| Yoruba | 0.95 (0.65-1.39) | 0.796 | 1.18 (0.76-1.84) | 0.464 |
| Igbo/Ibo | 1.46 (0.96-2.23) | 0.078 | 1.69 (1.03-2.78) | 0.037 |
| Northern minority | 1.36 (1.01-1.83) | 0.040 | 1.45 (1.04-2.04) | 0.030 |
| Southern minority | 1.44 (0.94-2.21) | 0.091 | 1.30 (0.79-2.15) | 0.296 |
| Unclassified | 1.00 (0.49-2.04) | 0.995 | 1.11 (0.50-2.51) | 0.792 |
| Current contraceptive method: Not using (RC) | Not included | - | 1.00 |  |
| Modern method | - | - | 0.07 (0.05-0.10) | <0.001 |
| Traditional method | - | - | 0.04 (0.02-0.09) | <0.001 |
| Working status: No (RC) | 1.00 |  | 1.00 |  |
| Yes | 0.81 (0.63-1.04) | 0.099 | 0.86 (0.66-1.13) | 0.284 |

Source: Computed from the 2024 NDHS Women Recode dataset. Notes: Outcome is coded 1 = infecund/menopausal and 0 = fecund. Both models adjust for residence, education, wealth quintile, union status, children ever born, religion, ethnicity and working status. Model 2 additionally includes the current contraceptive method. AOR = adjusted odds ratio; CI = confidence interval; RC = reference category.

The current contraceptive method showed the largest adjusted association in Model 2. Compared with women not using a method, modern-method and traditional-method users had markedly lower odds of an infecund/menopausal classification. This finding is treated as a measurement-proximal contemporaneous association rather than as evidence that contraceptive use alters the biological timing of menopause.

## Discussion

This study demonstrates that Nigerian women aged 45-49 are not a homogeneous group with respect to reproductive status. Although the infecund/menopausal classification was the predominant DHS exposure category, more than two in five women remained classified as fecund, and small proportions were pregnant or postpartum amenorrhoeic. This reinforces the need to distinguish late reproductive age from confirmed menopause. The outcome examined is a DHS demographic exposure classification, whereas clinical menopause is defined retrospectively after 12 consecutive months without menstruation, with no other physiological or pathological cause [1,10].

The bivariate distributions indicated socioeconomic patterning: the infecund/menopausal classification was more common among rural, less educated, poorer, and nonworking women. In Model 1, selected associations with education and wealth remained after adjustment. These patterns are broadly consistent with the literature linking socioeconomic disadvantage to earlier reproductive ageing or menopausal timing [12,13]. However, their attenuation in the contraceptive-method sensitivity model means they should be interpreted conservatively. The results provide evidence of social patterning, not proof that education or household wealth produces an earlier biological menopause.

Union status was the most consistent correlate. Women who were currently married or cohabiting, particularly those formerly married, had substantially higher adjusted odds of DHS-defined infecund/menopausal classification than women never in a union. Within a life-course interpretation, this finding points to differences in cumulative marital and reproductive trajectories rather than a biological effect of marital status. Earlier Nigerian work has shown that attainment of menopause varies across women’s demographic and reproductive profiles [14,15]. The present analysis adds national evidence that union history remains relevant when reproductive status is measured using the DHS demographic classification.

Ethnic variation was present but specification-sensitive. Northern minority ethnicity was associated with higher odds of an infecund/menopausal classification in both adjusted specifications, whereas the association for Igbo/Ibo women emerged only in Model 2. These differences should not be interpreted as fixed biological group effects. Ethnicity may reflect differences in union histories, fertility regimes, contraceptive practices, material circumstances and other social contexts not fully captured in this analysis. Nigerian menopause-related research has also highlighted the importance of contextual and relational circumstances in women’s experiences of later reproductive life [16].

The inclusion of the current contraceptive method materially altered the model. Users of modern and traditional methods had markedly lower odds of an infecund/menopausal classification than non-users. This is substantively plausible, as contraceptive use in women’s forties can signal continuing or perceived exposure to pregnancy risk [7,19]. Nonetheless, the variable was recorded contemporaneously with V623 and may partly reflect the same reproductive-exposure logic. The resulting odds ratios should therefore be interpreted as proximal associations rather than as evidence that contraceptive use delays or prevents menopause.

Children ever born highlighted the importance of model sensitivity. There was no bivariate difference in classification across parity categories, whereas women with five or more children had lower odds only in Model 1. The association disappeared after the contraceptive method was added. This finding is consistent with evidence that reproductive factors and natural menopause have complex, context-dependent relationships [20]. It also indicates that parity should not be prioritised over more stable markers, particularly union history, when interpreting the present results.

## Strengths and limitations

The study has three strengths. First, it uses nationally representative survey data to focus on women aged 45-49, an age group often subsumed within broader reproductive-age analyses. Second, the analysis explicitly preserves the distinction between DHS-defined infecund/menopausal status and clinically confirmed menopause. Third, the two-model strategy shows how findings change when a contemporaneously measured, measurement-proximal variable is introduced.

Several limitations constrain the scope of interpretation. V623 is a demographic exposure-status category, not a clinical measure of natural menopause or infertility. The cross-sectional design precludes temporal or causal inference. Residual confounding is possible because the NDHS does not include all clinical, behavioural or earlier-life determinants of reproductive ageing. The current contraceptive method is measured contemporaneously and substantially alters adjusted estimates. Finally, the never-in-union reference group is small, so the magnitude of union-status odds ratios should be interpreted cautiously, even though the direction is consistent across specifications.

## Implications for policy and research

Reproductive health services for women in their late forties should not assume universal menopause. Counselling on pregnancy risk, contraception and reproductive transition should recognise heterogeneity in exposure status. Population research and programme monitoring should also maintain a clear distinction between demographic classifications of infecund/menopausal status and clinically confirmed menopause. Future studies should combine nationally representative data with longitudinal, qualitative or clinically verified measures to clarify how marital trajectories, social inequalities and reproductive histories shape women’s transition out of fecundity in Nigeria.

## Conclusion

Among Nigerian women aged 45-49, DHS-defined infecund/menopausal status was the most prevalent but not universal, with a substantial proportion remaining classified as fecund. Union status was the most stable correlate across adjusted specifications, whereas education, wealth, parity and ethnic associations were more sensitive to whether the current contraceptive method was included. The findings support a life-course and social-determinants interpretation of late-reproductive-life exposure status, but they do not establish clinically confirmed menopause or causal determinants of reproductive ageing. A more precise policy and research response requires recognising that women nearing age 50 remain heterogeneous in their reproductive exposure and health-service needs.

## Data availability statement

The dataset used in this analysis is available from The DHS Program, following registration and approval to access the 2024 Nigeria Demographic and Health Survey Women Recode file.

## Notes

### Competing Interest Statement

The authors have declared no competing interest.

### Funding Statement

The author(s) received no specific funding for this work.

### Author Declarations

The study used de-identified secondary data from the NDHS. The DHS Program and the survey implementing agencies obtained informed consent from respondents during data collection. No respondent-identifying information was used in this analysis.

